# Use of Sociodemographic Information in Clinical Vignettes of Multiple-Choice Questions for Preclinical Medical Students

**DOI:** 10.1101/2022.11.09.22282150

**Authors:** Kelly Carey-Ewend, Amir Feinberg, Alexis Flen, Clark Williamson, Carmen Gutierrez, Samuel Cykert, Gary L. Beck Dallaghan, Kurt O. Gilliland

## Abstract

**Purpose:** To characterize the use of demographic data in multiple-choice questions from a commercial preclinical question bank and determine if there is appropriate use of different distractors.

**Background:** Multiple-choice questions for medical students often include vignettes describing a patient’s presentation to help guide students to a diagnosis, but overall patterns of usage between different types of nonmedical patient information in question stems has yet to be determined.

**Methods:** 380 of 453 randomly selected questions were included for analysis after determining they contained a clinical vignette and required a diagnosis. The vignettes and following explanations were then examined for the presence/absence of 11 types of demographic information, including age, sex/gender, and socioeconomic status. We compared both the usage frequency and relevance between the 11 information types.

**Results:** Most information types were present in less than 10% of clinical vignettes, but age and sex/gender were present in over 95% of question stems. Over 50% of questions included irrelevant information about age and sex/gender, but 75% of questions did not include any irrelevant information of other types. Patient weight and environmental exposures were significantly more likely to be relevant than age or sex/gender.

**Discussion:** Students using the questions in this study will frequently gain practice incorporating age and sex/gender into their clinical reasoning while receiving little exposure to other demographic information. Based on our findings, we posit that questions could include more irrelevant information, outside age and sex/gender, to better approximate real clinical scenarios and ensure students do not overvalue certain demographic data.

## Introduction

Multiple-choice questions (MCQs) are commonly employed in preclinical medical curricula for both review of material and assessment of competency. Examinations in medical school are often made up entirely of MCQs, and each of the licensing exams through the United State Medical Licensing Examination (USMLE) three-step program are comprised largely of multiple-choice items. MCQs often rely on the use of clinical vignettes (a description of a patient, including information about their symptoms, medical history, and/or sociodemographic context) as a key part of the question structure. This style of questioning can be referred to as case-based, which attempts to imitate real-life clinical scenarios while also requiring higher-order cognitive processing from the students [1]. The information included in a vignette should allow a student reading the question to differentiate between the correct answer option and the remaining incorrect options [2].

However, not all information found in a clinical vignette is relevant to determining the correct answer option. Current guidelines for question-writing employed by the National Board of Medical Examiners (NBME) advise that clinical vignettes feature patient characteristics that “are clinically relevant and/or aid in distractor quality” as well as characteristics that “add to the overall exam-level representation of the referenced patient population” [2]. These guidelines lead question writers to reach a balance of information relevancy. If a clinical vignette includes too much irrelevant information about a patient, then the question may become confusing and cumbersome to students’ progression through timed exams. Conversely, if a question features only information that directly informs the exact diagnosis, the question no longer imitates real clinical scenarios.

The use of demographic information has been a point of notable scrutiny in recent years. Demographic information, especially race, has been employed inconsistently in ways that can create false associations and affirm biases amongst students. In a survey of 22 medical students by Mosley et al., all participants reported viewing race in clinical vignettes as a “clue” towards the correct option while also describing that they saw mentions of non-white races as more likely to be relevant [3]. This finding was affirmed by Ripp et al. who, in an analysis of a Step 1 USMLE question bank, found that race was more likely to be relevant if the patient’s race was African American vs. Caucasian, and these relevant uses of race were more often employed as a proxy for genetics rather than to describe the patient’s sociocultural context, despite race being a social construct with little biological relevance [4]. These tendencies in overall inclusion and relevancy can lead students to develop powerful associations between specific patient populations and certain illnesses that, while helpful on examinations which rely on these associations, may ultimately affirm bias and impede students’ ability to determine information relevancy in real clinical situations.

In practice, clinicians immediately perceive certain patient characteristics upon meeting them (such as age, sex, race, and weight), though this visual approximation is not always accurate [5]. Other types of demographic information, like occupation or sexual orientation, might only be revealed by specific history-taking. Regardless, each of these demographic factors carry cultural stigma and societal significance that students must learn to navigate in order to develop structural competency, a skillset focused on attention to factors that influence health outcomes above the level of individual interactions [6].

The specific usage of race in clinical vignettes has been examined in-depth in some MCQ banks for preclinical medical students, but overall patterns of demographic information inclusion and usage have not been systematically evaluated. The ability of students to learn structural competency regarding patient sociodemographics from MCQ banks depends on the inclusion frequency of these demographic variables and the subsequent relevancy to those questions. As such, the authors of this study sought to quantify the inclusion and relevance of 11 types of demographic information in the clinical vignettes of a MCQ bank for preclinical medical students.

## Methods

### Question analysis

The USMLE-Rx Qmax Step 1 question bank from *Scholar-Rx* was identified as a widely used educational resource for preclinical medical students that is frequently employed in medical school curriculum development; among others, the University of North Carolina School of Medicine provides this resource to its student body and mandates its usage. The question bank is comprised of 2380 MCQs developed using NBME question-writing guidelines to help students prepare for the United States Medical Licensing Examination Step 1. Verbal authorization to employ this question bank was given by the company’s Vice President for Medical Education. MCQs (or “items”) were sampled randomly from this bank and subsequently analyzed.

Items follow a typical format of a question stem (comprised of a clinical vignette followed by a lead-in/question) and answer options (typically 4-8, of which only one is correct). After choosing an option, whether right or wrong, the student is given an explanation. The explanation is roughly one page of text interpreting the vignette, providing background about the content of the item, and detailing why the correct option was correct and the incorrect options were incorrect.

For an item to address the aims of this study, it needed to include a clinical vignette in the stem and require the student to make a diagnosis in order to select the correct option. Of 453 items randomly sampled from the item bank, 380 were included for analysis after determining they met both criteria. Both the vignettes and explanations of these items were individually examined for the presence or absence of 11 types of demographic information: age, race/ethnicity, sex/gender, sexual behaviors/orientation, environmental exposures, socioeconomic status (SES), illicit drug use, alcohol use, nicotine use, diet, and weight/body mass index (BMI). An information type included in the explanation would not be counted as present if it was not also present in that question’s vignette, as this information came after choosing an option and could not inform decision-making. Over the course of question analysis, terms associated with each category were compiled into a data dictionary for consistency (**Supplementary Data 1**). Each question was analyzed by one of four authors, with the authors meeting regularly during coding to cross-check data, present updates to the data dictionary, and discuss discrepancies or complexity in the coding scheme.

Once the presence/absence of the information types was determined for each clinical vignette and matched explanation, the whole set was analyzed for prevalence and relevance of each information type. The number of irrelevant information types in each item was also determined.

The prevalence of an information type in item stems refers to the proportion of all sampled items that included a specific type of patient information in the clinical vignette. The relevance of an information type refers to the proportion of item stems with that information type in the vignette that also cited the information type in its explanation as a justification for the correct option. For example, if 40 questions out of 380 explicitly mentioned the patient’s socioeconomic status in the vignette, and 5 of these 40 questions cited the patient’s SES in the explanation as support for choosing the correct answer option, then the “SES” information type would have a prevalence of 10.5% (40 out of 380) and a relevance of 12.5% (5 out of 40).

The number of irrelevant information types per item was determined by subtracting the number of relevant information types in an item’s explanation from the total number of information types in that item’s clinical vignette, yielding the number of information types included in the vignette that were not cited as important in determining the correct answer option.

### Statistical analysis

Prevalence proportions were compared using McNemar’s test for equality of matched proportions on all 55 pairwise comparisons with a Bonferroni adjustment [7,8]. Relevance proportions were compared using Fisher’s exact test for equality of two unmatched sample proportions on all 55 pairwise comparisons with a Bonferroni adjustment. The difference in irrelevant information types per question when counting and not counting age and sex/gender was tested using a large-sample normal-approximation z-test. Large-sample McNemar’s tests, Fisher’s exact tests, and z-tests were performed in Microsoft Excel (**Supplementary Data 1**) and exact McNemar’s tests were performed in R version 4.1.3 (R Core Team, Vienna, Austria) using the package “exact2×2” version 1.6.6 (**Supplementary Data 2**).

## Results

The data shows a major discrepancy in prevalence of sex/gender and age when compared to the other information types examined. While most information types were present in less than 10% of clinical vignettes, age and sex/gender were present in over 95% of question stems, significantly higher than all other types examined (p-values: <0.001) (**Figure 1**). Age was found in 374 (98.4%) vignettes and sex/gender was included in 371 (97.6%) vignettes. The next most prevalent information types examined were the broad environmental exposure category, which was present in 57 (15.0%) clinical vignettes, and weight/BMI, which was present in 45 (11.8%) vignettes. Environmental exposures and weight/BMI were significantly more prevalent than the remaining types, other than nicotine use, alcohol use, and each other (p-values: <0.05) **(Table 1**).

**Table 1:** Raw data for prevalence and relevance (counts and proportions) of each information type. Prevalence refers to the count and proportion of all 380 analyzed items that included the info type in the vignette. Relevance refers to the count and proportion of items with the info type in the vignette that also cited the information as relevant in the explanation (A). P-values are shown for all comparisons of each information type’s prevalence proportion to all others by McNemar’s test (B). P-values are also shown for all comparisons of each information type’s relevance proportion to all others using Fisher’s exact test (C).

| A. Raw Data | Age | Race/<br>Eth. | Sex/<br>Gender | Sex.<br>Behavior | Env.<br>Exp. | SES | Ill.<br>Drug<br>Usage | Alc.<br>Use | Nic.<br>Use | Diet | BMI |
| --- | --- | --- | --- | --- | --- | --- | --- | --- | --- | --- | --- |
| Prevalence<br>Proportion | 0.98 | 0.06 | 0.98 | 0.03 | 0.15 | 0.02 | 0.04 | 0.07 | 0.08 | 0.02 | 0.12 |
| Prevalence<br>Count | 374 | 23 | 371 | 12 | 57 | 6 | 17 | 27 | 30 | 7 | 45 |
| Relevance<br>Proportion | 0.26 | 0.22 | 0.17 | 0.42 | 0.53 | 0.33 | 0.41 | 0.22 | 0.33 | 0.43 | 0.58 |
| Relevance<br>Count | 97 | 5 | 64 | 5 | 30 | 2 | 7 | 6 | 10 | 3 | 26 |

| B. P-value for prevalence comparisons | Age | Race/ Eth. | Sex/ Gender | Sex. Behavior | Env. Exp. | SES | Ill. Drug Usage | Alc. Use | Nic. Use | Diet | BMI |
| --- | --- | --- | --- | --- | --- | --- | --- | --- | --- | --- | --- |
| Age |  |  |  |  |  |  |  |  |  |  |  |
| Race/ Eth. | <0.01 |  |  |  |  |  |  |  |  |  |  |
| Sex/ Gender | >0.99 | <0.01 |  |  |  |  |  |  |  |  |  |
| Sex. Behavior | <0.01 | >0.99 | <0.01 |  |  |  |  |  |  |  |  |
| Env. Exp. | <0.01 | <0.01 | <0.01 | <0.01 |  |  |  |  |  |  |  |
| SES | <0.01 | 0.16 | <0.01 | >0.99 | <0.01 |  |  |  |  |  |  |
| Ill. Drug Usage | <0.01 | >0.99 | <0.01 | >0.99 | <0.01 | >0.99 |  |  |  |  |  |
| Alc. Use | <0.01 | >0.99 | <0.01 | 0.29 | 0.04 | <0.01 | >0.99 |  |  |  |  |
| Nic. Use | <0.01 | >0.99 | <0.01 | 0.11 | 0.11 | <0.01 | 0.99 | >0.99 |  |  |  |
| Diet | <0.01 | 0.34 | <0.01 | >0.99 | <0.01 | >0.99 | >0.99 | 0.06 | <0.01 |  |  |
| BMI | <0.01 | 0.22 | <0.01 | <0.01 | >0.99 | <0.01 | <0.01 | 0.97 | >0.99 | <0.01 |  |

**Table 1:** Raw data for prevalence and relevance (counts and proportions) of each information type.
| C. P-value for relevance comparisons | Age | Race/ Eth. | Sex/ Gender | Sex. Behavior | Env. Exp. | SES | Ill. Drug Usage | Alc. Use | Nic. Use | Diet | BMI |
| --- | --- | --- | --- | --- | --- | --- | --- | --- | --- | --- | --- |
| Age |  |  |  |  |  |  |  |  |  |  |  |
| Race/ Eth. | >0.99 |  |  |  |  |  |  |  |  |  |  |
| Sex/ Gender | >0.99 | >0.99 |  |  |  |  |  |  |  |  |  |
| Sex. Behavior | >0.99 | >0.99 | >0.99 |  |  |  |  |  |  |  |  |
| Env. Exp. | <0.01 | 0.57 | <0.01 | >0.99 |  |  |  |  |  |  |  |
| SES | >0.99 | >0.99 | >0.99 | >0.99 | >0.99 |  |  |  |  |  |  |
| Ill. Drug Usage | >0.99 | >0.99 | >0.99 | >0.99 | >0.99 | >0.99 |  |  |  |  |  |
| Alc. Use | >0.99 | >0.99 | >0.99 | >0.99 | >0.99 | >0.99 | >0.99 |  |  |  |  |
| Nic. Use | >0.99 | >0.99 | >0.99 | >0.99 | >0.99 | >0.99 | >0.99 | >0.99 |  |  |  |
| Diet | >0.99 | >0.99 | >0.99 | >0.99 | >0.99 | >0.99 | >0.99 | >0.99 | >0.99 |  |  |
| BMI | <0.01 | 0.25 | <0.01 | >0.99 | >0.99 | >0.99 | >0.99 | 0.17 | >0.99 | >0.99 |  |

*Abbreviations: Race/ Eth. = Race/Ethnicity; Sex. Behavior = Sexual Behaviors and Orientation; Env. Exp. = Environmental Exposures; SES = Socioeconomic Status; Ill. Drug Usage = Illicit Drug Usage; Alc. Use = Alcohol Use; Nic. Use = Nicotine Use; BMI = Weight and Body Mass Index*

**Figure 1:**
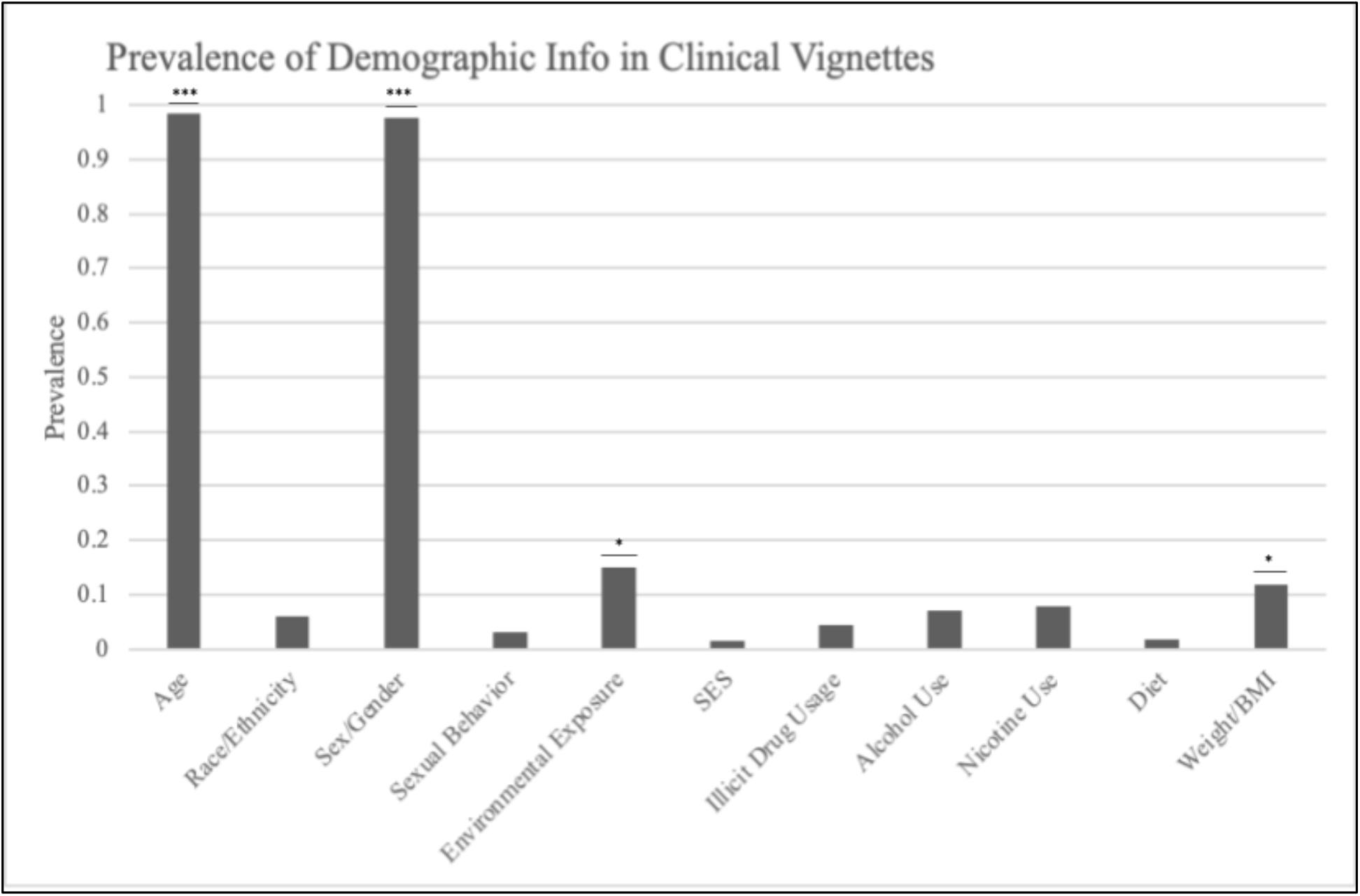
Prevalence of each demographic information type across all questions analyzed. Reflects the proportion of all 380 analyzed questions that included the specified information type in its clinical vignette. Pairwise comparisons were performed using a McNemar’s tests for matched proportions with a Bonferroni adjustment. Age and sex/gender were significantly more prevalent than all other information types except each other (p-values: <0.001). Of the remaining types, the prevalence of environmental exposure was significantly greater than those remaining except for weight/BMI and nicotine use (p-values: <0.05). Weight/BMI was also significantly more prevalent than the remaining information types other than environmental exposures, alcohol use, and nicotine use (p-values: <0.01).

While prevalence was highly disparate between some of these information types, their relevance to selecting the correct answer option in each question was more even. Sex/gender was least relevant, being cited in each question’s explanation as useful in making the correct diagnosis in only 64 of 371 questions (17.3%) (**Figure 2**). Race/ethnicity, alcohol use, and age showed similar relevance at 21.7%, 22.2%, and 26.0%, respectively. Conversely, diet, environmental exposures, and weight/BMI were most frequently relevant, cited in the explanation 42.8%, 52.6%, and 57.8% of the time, respectively. Environmental exposure was significantly more likely to be relevant than sex/gender and age, as was weight/BMI (p-values: <0.01) (**Table 1**). Of the 5 questions that cited race as relevant to selecting the correct answer choice, 4 employed races as a demonstrator of increased genetic and/or heritable risk of certain conditions (sickle cell disease, cystic fibrosis, etc.) and none of them discussed race in terms of sociopolitical factors, economic factors, or structural access to health care.

**Figure 2:**
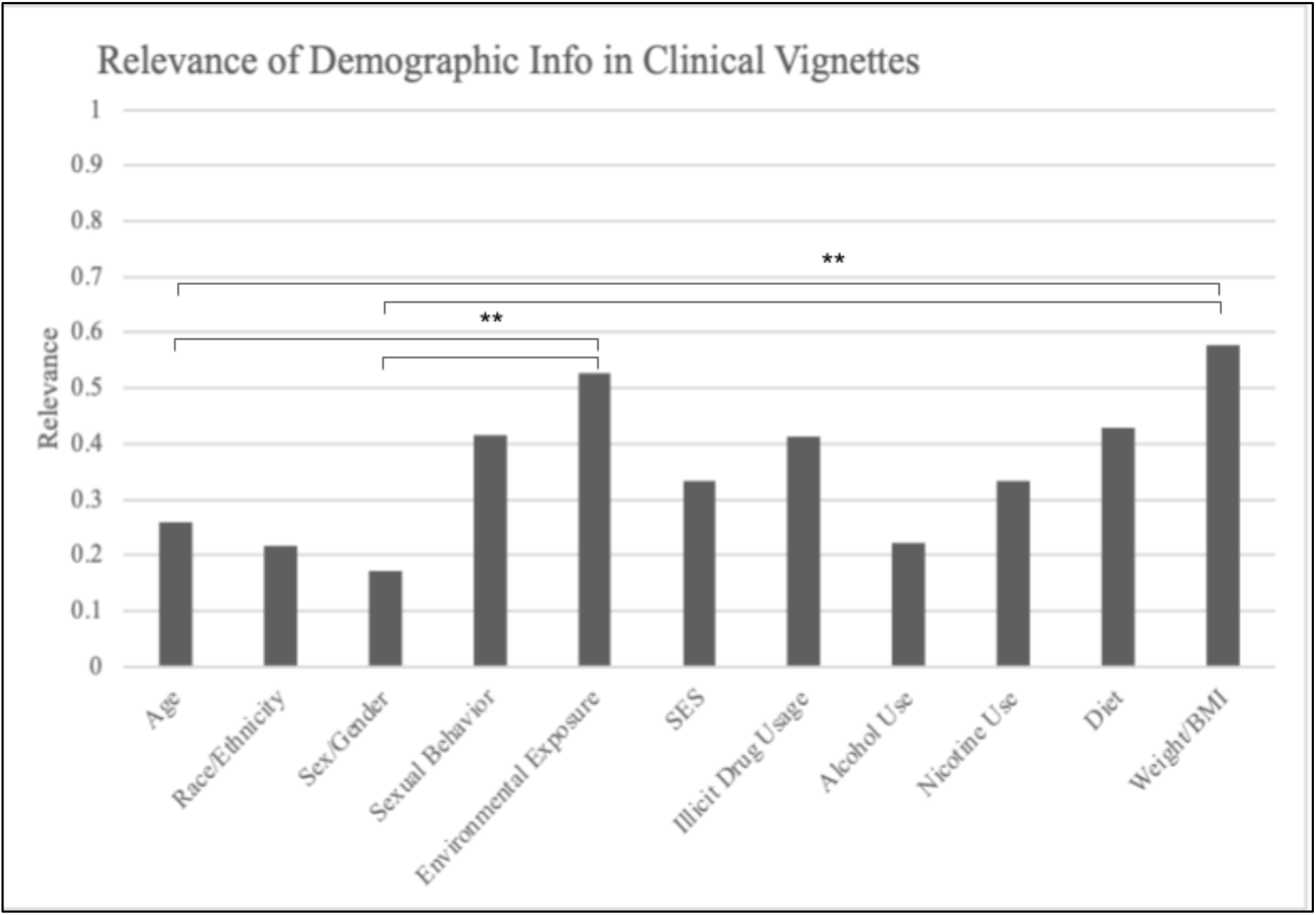
Relevance of information types in clinical vignettes. Refers to the proportion of questions with a given information type in the vignette that also cited the information as relevant in the explanation. Proportions were compared to each other using Fisher’s exact test with a Bonferroni adjustment. Relevances of both environmental exposure and weight/BMI were significantly higher than both age and sex/gender (p-values <0.01).

The presence of irrelevant information in each clinical vignette was also examined, with irrelevant information being information found in the vignette but not cited in the explanation. The average question that met analysis criteria contained 1.87 irrelevant information types in its clinical vignette (**Figure 3A**). When age and sex were excluded from this count, the number of irrelevant non-age/sex/gender information types in each question was 0.3 on average (**Figure 3B**). Over 50% of questions included irrelevant information about age and sex/gender. However, outside of age and sex/gender, 75% of questions did not include any irrelevant demographic information of any other type examined.

**Figure 3:**
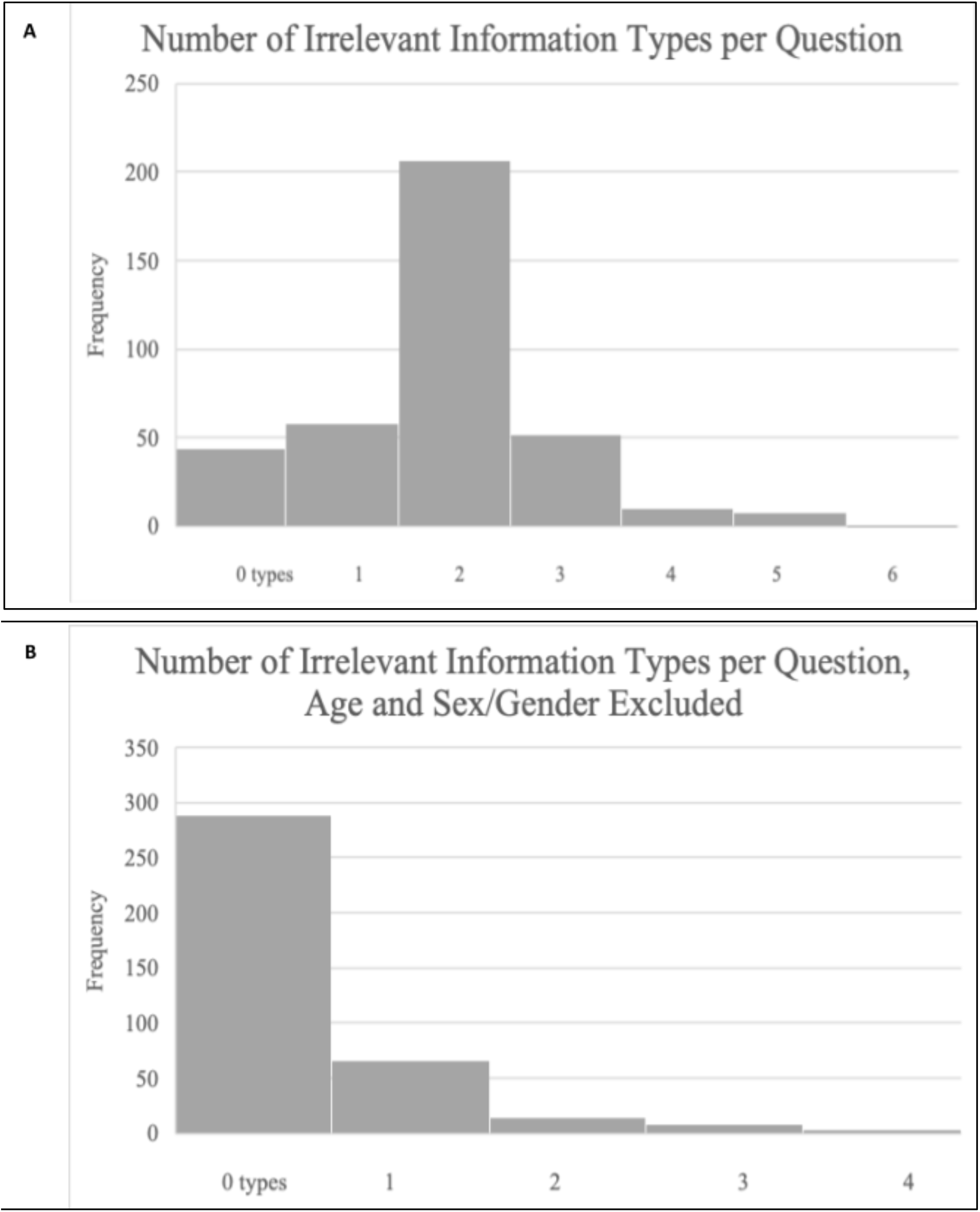
Number of irrelevant information types included in clinical vignettes. The average number of irrelevant information types per question was 1.87 (A). The same calculation, when excluding age and sex/gender, yields an average of 0.34 irrelevant information types per question (B). The difference between these two values (1.54) is statistically significant (p-value: <0.001).

## Discussion

The findings of this study demonstrate a few key inconsistencies in the use of demographic information in the clinical vignettes examined. Students using these questions will receive exposure to age and sex information about their patients in almost every question and will be presented with these characteristics more commonly in the first sentence of the vignette, while other information types tend to be evenly dispersed throughout the vignette. These two types are certainly clinically relevant, as sex generally carries information about the organs present in a person while many conditions are risk stratified by age. They are also the two to which clinicians would most frequently have access given the common inclusion and prominent display of age and sex assigned at birth on health records. They were also by far the most commonly included irrelevant information of any of those examined, comprising 82% of all superfluous sociodemographic information in the clinical vignettes.

While it can be supported that these two variables are the most prominent in the MCQs studied, the magnitude of the discrepancy between these factors and the others examined warrants examination. Students using USMLE-Rx will be exposed to age and sex of their patients in almost every item (compared to, at most, 15% for any other demographic variables) and will have to incorporate age into their clinical reasoning in 25.5% of all questions while recognizing the information as irrelevant in 72.8% of all questions. The ability to collect sociodemographic information and determine its relevancy is important in clinical care as, again, thorough patient history will often yield information about sex, age, race, socioeconomic status, diet, drug usage, etc. even when this information does not directly inform a patient’s condition. In the questions studied, students are able to practice using age and sex in almost every item while receiving little exposure to the other information types. As mentioned before, visual categorization of people on the basis of perceived age, sex, gender, race, and weight, among other things, often occurs unintentionally and automatically, though not always correctly [5]. The clinical vignettes in this study imitated the frequency with which clinicians must evaluate and work with/around age and sex information while almost never including other types of sociodemographic information examined.

One possible solution to address these inconsistencies would be to adjust the irrelevant usage of more represented demographic types (age, sex/gender) and less represented demographic types (socioeconomic status, race, sexual behaviors/orientation, etc.) to reach a more balanced distribution. As it stands, over 75% of clinical vignettes include irrelevant information about age or sex/gender while less than one in three include irrelevant information of any other type. A reduction in age and sex inclusion combined with an increase in use of other information types would maintain question length while increasing the breadth and overall representation of patient sociodemographic information across clinical vignettes. This would aid in the NBME “Constructing Written Test Questions for the Health Sciences” Guide’s stated goal of “add[ing] to the overall exam-level representativeness of the referenced patient population” without changing the degree to which the information included in each vignette “[is] clinically relevant and/or aids in distractor quality.” [2]

Special attention must be paid to some of these variables when discussing their inclusion due to their previously documented history of misuse and the danger of perpetuating bias [4]. Race, for example, is a social construct with a long history of associated cultural stigma, and its usage in many clinical vignettes has erroneously relied on race as a proxy for genetic relation. This can be dangerous for rising clinicians, as Hoffman et al. established that the incorrect conception of race as having a biological and genetic basis was correlated with racially biased pain assessment [10]. As such, many previous authors on the topic of race in medical education materials have argued for the overall omission of race from clinical vignettes to prevent perpetuation of stereotypes [4, 11]. However, the current inclusion of race in only 6% of question stems, when compared with age and sex being present in over 95%, does not reflect the frequency with which clinicians are presented with these types of information. Additionally, the overall lack of race inclusion in questions may contribute to the findings that students overvalue race when featured in vignettes, especially when referring to Black patients [3]. Randomized inclusion of irrelevant demographic information with the goal of matching overall population proportions could address these concerns while relying on the randomization process to prevent perpetuation of bias on a broad scale.

Finally, non-randomized employment of these less-prevalent information types could be used to actively assess and teach students about their impacts on health outcomes in the United States. While race is a poor correlate for genetics and biology, its political relevance and connection with access to health infrastructure is well-documented and a key part of “structural competency”, as described by Metzl et al. and Tsai et al. [6, 9]. However, in the small number of questions examined in this study that employed race to guide students to the correct option, none of them referenced the social, political, or economic connections between race and health outcomes. The small number of relevant uses of race should be shifted to focus on its social basis rather than biological presumptions. Additionally, socially significant demographic data in questions could be employed to educate students about the social context of disease in certain patient populations, address preexisting biases, and improve the population-level representativeness of the question bank.

While the major conclusions drawn from these results should be unaffected by small errors in data-coding, the basis of this study is fundamentally qualitative, and some limitations should be noted. The nuance of certain categorizations (e.g., sex versus gender, race versus ethnicity, whether immigrant status carries racial information) were deemed too complex and rarely employed in the question bank to warrant distinct categorization. Another important limitation that arose anecdotally involves situations in which the authors felt that a piece of demographic information in a vignette was employed in a relevant way, but the explanation of the question did not explicitly cite it as relevant. While this occasional loss of relevance may have impacted the overall power of our analyses, choosing to include only explicit statements of relevance helped to reduce the impact of author bias on interpretation of these questions. Finally, question bank used for this study was developed using NBME question-writing guidelines and follows the structure of USMLE Licensing exams, but it may not be representative of USMLE exam content or of other education resources intended for the same student population. Therefore, while our analysis sheds light on the output of current MCQ-writing practices, our exact findings may not apply to other question banks.

Future studies in this area could apply similar methodologies to other preclinical or clinical medical educational resources. Additionally, more focused analysis on the exact usage of sex and gender or different racial and ethnic groups could offer a more detailed view on how these demographics are employed in MCQs. Finally, a similar investigation into how frequently practicing clinicians encounter and use each of the above demographic information types could provide a scaffold for question-writers to ensure that their use of demographic information in clinical vignettes accurately reflects their importance in actual clinical settings.

## Supporting information

Supplementary Data 2, R script as text file

## Data Availability

All data produced in the present study are available upon reasonable request to the authors

## Structured Disclosures

## Acknowledgements

We thank the UNC Offices of Medical Student Education for their support, and to the *Scholar-Rx* corporation for use of their question bank.

## Ethical Approval

Not applicable.

## Disclaimers

None.

## Previous Presentations

None.

## Legends

**Supplementary Data 1: Excel spreadsheet used for data collection and analysis**. Tabs show the final coding dictionary, data collected for each analyzed item, analysis to compare prevalence proportions between different information types, and analysis to compare relevance proportions between difference information types.

**Supplementary Data 2: R script used to compare prevalence proportions when sample size was small**. The script was run on R version 4.1.3 using the package “exact2×2” version 1.6.6.

